# Increased luminal area of large conducting airways in patients with COVID-19 and post-acute sequelae of COVID-19 A retrospective case-control study

**DOI:** 10.1101/2024.02.29.24303556

**Authors:** Solomiia Zaremba, Alex J. Miller, Erik A. Ovrom, Jonathon W. Senefeld, Chad C. Wiggins, Paolo B. Dominelli, Ravindra Ganesh, Ryan T. Hurt, Brian J. Bartholmai, Brian T. Welch, Juan G. Ripoll, Michael J. Joyner, Andrew H. Ramsook

## Abstract

**Background:** Coronavirus disease 2019 (COVID-19) is associated with enlarged luminal areas of large conducting airways. In 10-30% of patients with acute COVID-19 infection, symptoms persist for more than 4 weeks (referred to as post-acute sequelae of COVID-19, or PASC), and it is unknown if airway changes are associated with this persistence. Thus, we aim to investigate if luminal area of large conducting airways is different between PASC and COVID-19 patients, and healthy controls.

**Methods:** In this retrospective case-control study seventy-five patients with PASC (48 females) were age-, height-, and sex-matched to 75 individuals with COVID-19 and 75 healthy controls. Using three-dimensional digital reconstruction from computed tomography imaging, we measured luminal areas of seven conducting airways, including trachea, right and left main bronchi, bronchus intermediate, right and left upper lobe, and left lower lobe bronchi.

**Findings:** Airway luminal areas between COVID-19 and PASC groups were not different (p>0.66). There were no group differences in airway luminal area (PASC vs control) for trachea and right main bronchus. However, in the remaining five airways, airway luminal areas were 12% to 39% larger among PASC patients compared to controls (p<0.05).

**Interpretation:** Patients diagnosed with COVID-19 and PASC have greater airway luminal area in most large conducting airways compared to healthy controls. No differences in luminal area between patients with COVID-19 and PASC suggest persistence of changes or insufficient time for complete reversal of changes.

**Funding:** National Heart, Lung, and Blood Institute (F32HL154320 to JWS; 5R35HL139854 to MJJ); Postdoctoral Fellowship from the Natural Sciences and Engineering Research Council of Canada (AHR).

## Introduction

COVID-19 is a clinical syndrome caused by severe acute respiratory syndrome coronavirus 2 (SARS-CoV-2) primarily transmitted person to person via respiratory droplets.^1,2^ Patients may present with varying severity of infection: from asymptomatic to severe form with development of acute respiratory distress syndrome. In some cases, symptoms may persist for longer than four weeks, referred to as post-acute sequelae of SARS-CoV-2 infection (PASC) or long COVID.^3^ Reported incidence of PASC varies between 10 to 30% in nonhospitalized patients and about 50 to 70% in hospitalized patients.^4,5^ Manifestations of PASC include constitutional symptoms, and respiratory symptoms such as dyspnea on exertion, persistent cough, and shortness of breath.^6^ While many studies report on respiratory symptoms of COVID-19 and PASC, less is known about anatomical or pathophysiological changes in airways. Findings from studies have shown increase in diameter of the trachea in COVID-19 patients that is proportional to severity of the infection and might be associated with worse prognosis.^7^

Previously, our group has also evaluated large conducting airways in patients with recent COVID-19 infection by measuring airway luminal area from patients computed tomography (CT) imaging.^8^ CT has been widely used during the pandemic as a tool for patient diagnostics, evaluation, and management.^9,10^ Analyses showed that in patients with COVID-19 infection, who did not develop long term sequelae (PASC), COVID-19 was associated with increased large conducting airways luminal area compared to healthy controls. This finding provides evidence of a potential association of increased airway area with COVID-19 infection.

Taking into consideration persistence of symptoms after initial infection, our group has continued investigating possible large airway involvement in PASC pathogenesis in an attempt to determine if the association observed during acute phase subsides over time. Available data on airway pathological changes in COVID-19 survivors are inconclusive. Some studies report permanent pathologic dilation to the airways on CT imaging months after initial diagnosis,^6–9^ known as bronchiectasis, and others discuss reversibility of pathological airway changes.^11^

Accordingly, our study aimed to determine the relationship between large conducting airway size and PASC. This retrospective, case-control study used chest CT imaging to test the hypothesis that PASC is associated with enlarged airway luminal area in patients with PASC compared to their healthy matched counterparts. Our secondary exploratory aim was to determine if airway changes observed in patients with COVID-19 persist in patients with PASC or subside over time.

## Methods

### Ethical approval

The study was approved by Mayo Clinic Institutional Review Board (IRB 17– 008537) and was conducted in accordance with the ethical standards of the Declaration of Helsinki except for registration in a database. Informed consent was waived as no identifiers were used, and already existing data were extracted from patient electronic health records (EHR). The waiver was approved by Mayo Clinic Institutional Review Board.

### Participants

Adult patients (≥18 years old) diagnosed with PASC, who underwent computed tomography (CT) between January 2020 and August 2022 met the criteria for inclusion. PASC was defined as persistent or new symptoms in patients with COVID-19 more than four weeks after onset of the disease. In each case, diagnosis was confirmed by physician. Overall exclusion criteria were similar in all three groups. Graphical presentation of subject inclusion and exclusion procedure is represented in Figure 1.

**Figure 1.**
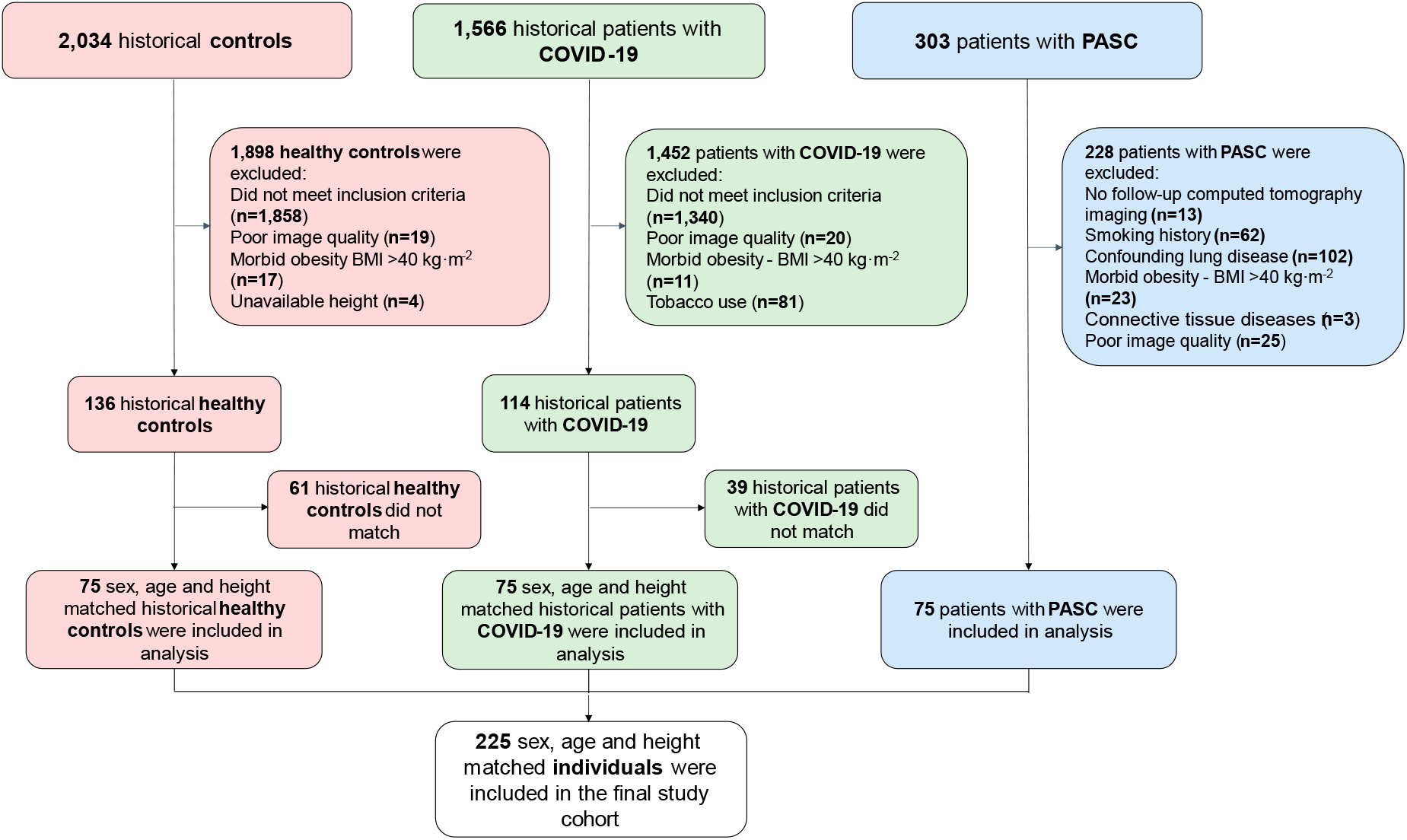
Flowchart of long COVID patient inclusion, exclusion and matching with the acute COVID-19 patients and healthy controls for the study. PASC, post-acute sequelae of COVID-19.

#### PASC

A cohort of 303 patients with PASC were considered for inclusion and underwent further screening of medical history in EHR. A total of 228 patients were excluded from this cohort. Among excluded, 102 individuals had pulmonary comorbidities (chronic obstructive pulmonary disease, asthma, cystic fibrosis, interstitial lung disease, pulmonary malignancy, history of pulmonary embolism, lung lymphangiomyomatosis, obstructive sleep apnea, history of congenital heart/lung disease). Another 13 patients were missing CT imaging in their EHR. Sixty-two excluded patients had smoking or tobacco use history, 23 had morbid obesity (body mass index ≥ 40 kg·m^-2^), and three patients were excluded due to connective tissue diseases (systemic lupus erythematosus and rheumatoid arthritis). Finally, 25 patients were excluded due to poor image quality during the data extraction process, as the software was not able to analyze imaging for further measurements.

#### COVID-19

The COVID-19 group included 1,566 historical patients from our previous study,^8^ with CT imaging performed between March 2020 and August 2021. The COVID-19 group was defined as patients in acute phase of COVID-19 or at early recovery, who did not experience any persistent COVID-19 symptoms. Exclusion criteria for this group were similar to those in PASC group, encompassing patients with comorbidities (n=1,340), poor image quality (n=20), morbid obesity with BMI ≥ 40 kg·m^-2^ (n=11), history of smoking or tobacco use (n=81). Patients that were unable to be matched to PASC patients (n=39) were excluded from the analysis.

#### Control

The control group initially included 2,034 subjects from our previous study,^8^ who underwent CT scanning due to suspected pulmonary embolism, that was not confirmed. The diagnostic imaging for these patients was conducted between March 2009 and March 2018 − before the pandemic. EHRs of potential controls were screened by an investigator and 1,898 subjects were excluded from the cohort. Sixty-one control subjects did not match PASC cohort and were subsequently excluded from the study.

Each compared group included 75 sex-, age-, and height-matched subjects. COVID-19 and PASC groups were stratified by sex, and two sequential nearest neighbor-matching algorithms were used to match patients based on both height (primary) and age (secondary). This algorithm was used in our previous study to match COVID-19 patients and healthy controls, and the same cohort was matched with PASC patients. Once participants in all groups were identified, CT images were analyzed for luminal area of the large conducting airways.

### Image acquisition

Standardized CT algorithms for routine thoracic imaging are utilized by our hospital and were broadly described before.^8,12-14^ Imaging was taken at end-inspiratory phase of breathing, although patients were not instructed to inhale until total lung capacity. A posterior-anterior and lateral topogram was obtained at 120 kV (with a standard milliampere-second value of 140) and 35 mA. Acquired images were then reconstructed at 1.5 mm and 3 mm slice thickness in the axial and coronal plane using a B46 kernel. Maximal intensity projections in both planes had a slice thickness of 10 mm and reconstruction increment of 2.5 mm.

### Airway luminal area measurement

We employed Aquarius Intuition software (AQi; Tera Recon, Foster City, CA, USA) to extract CT chest imaging from patient EHRs. This software performs three-dimensional reconstructions of the lungs of each patient along with the large conducting airways. AQi utilizes images in transverse, coronal, and sagittal planes to create a 3D model of large airways with clear differentiation between wall and lumen (Figure 2). Thus, investigators were able to measure luminal areas of the large conducting airways, including trachea, bronchus intermedius, right main and right upper lobe bronchi, left main bronchus, left upper and lower lobe bronchi. Measurements were taken at three points of each airway – proximal, middle, and distal. For the trachea, proximal point was defined as a point immediately below cricoid cartilage, and distal as a point just prior to trachea anatomical bifurcation. For bronchi, the proximal point was defined as a cross-sectional point directly below anatomical bifurcation of the airway of greater caliber, and distal as an airway cross-sectional point that preceded bronchial bifurcation. Middle of each airway was considered halfway between proximal and distal points.

**Figure 2.**
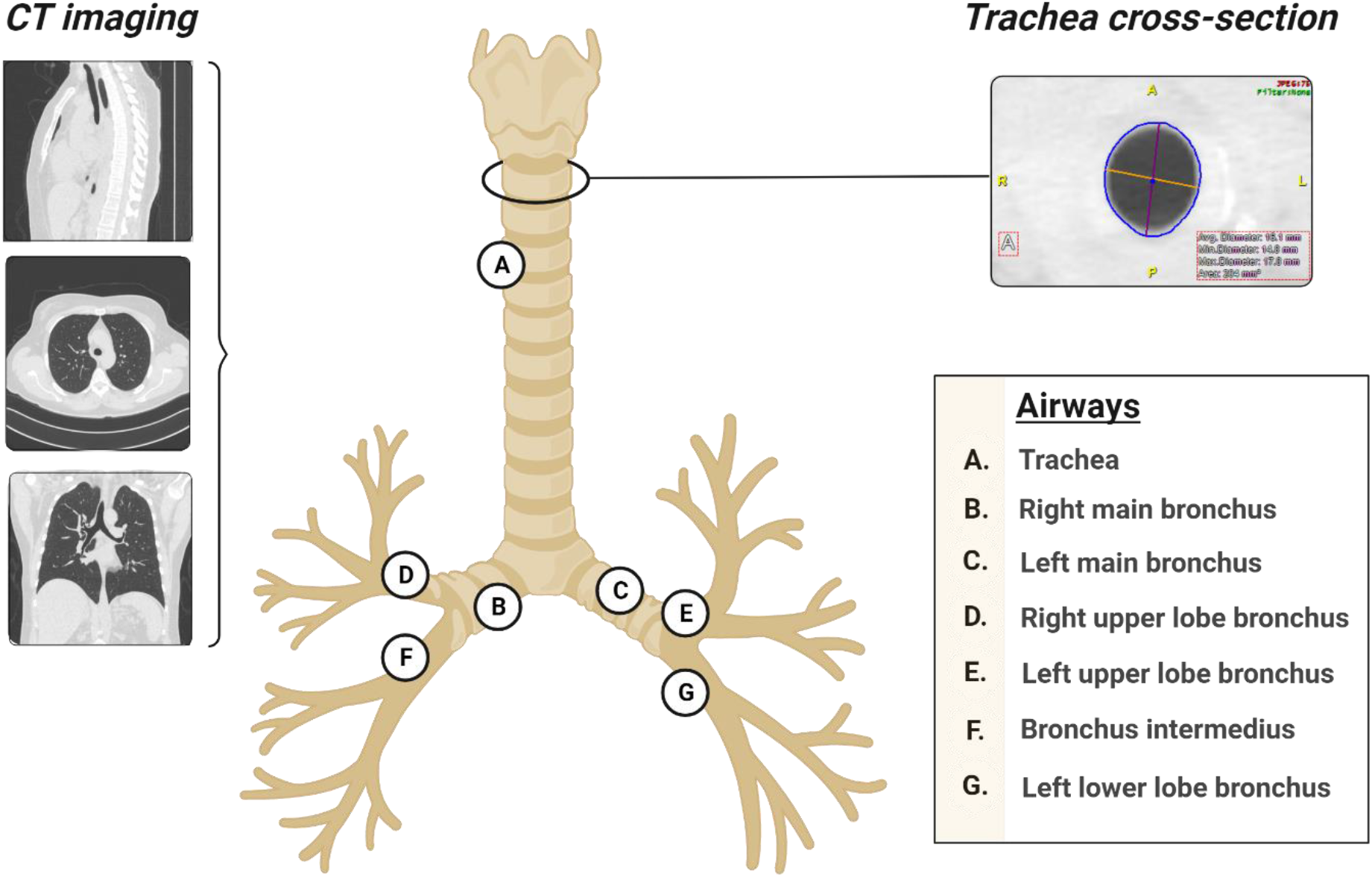
Aquarius Intuition (AQi) software of the Tera Recon company. Measurement of the trachea cross-sectional luminal area at proximal point. CT, computed tomography. Figure created with BioRender.com

### Statistical analysis

Assumptions of normality were contested with Shapiro-Wilk tests and assumptions of homoscedasticity were confirmed with Levene’s test. Shapiro-Wilk tests determined that demographic characteristics (age, height, weight, body mass index) and airway size data were not normally distributed. Thus, Kruskal-Wallis H test was used to compare demographic characteristics and airway luminal area measurements between the three groups (COVID-19, PASC, and control). Mann-Whitney U test was used to compare time between diagnosis and CT imaging date between two groups (COVID-19 and PASC). Multiple comparison tests were performed using the Dunn test and α values were adjusted using the Bonferroni method.

Statistical models were performed in duplicate using two representations of luminal airway size— the average of three measurements (proximal, middle, and distal points) and one measurement (middle point). Interpretation of findings largely did not differ between the statistical models, and findings using the average of three airway measurements are presented in the text. Results of analyses using the single measurement (middle point) are presented in supplemental information.

Descriptive statistics are presented as median (interquartile range) within the text, tables, and figures. Reported *p*-values are two-sided and the interpretation of findings was based on *p* < 0.05. Analyses were performed in IBM Statistical Product and Service Solutions (version 28, Armonk, New York, USA). Figures were created using GraphPad Prism software (version 9, La Jolla, California, USA).

## Results

### Demographics

Participant demographics are presented in Table 1. In accordance with the methodological design of the study, there were no group differences in sex, age, and height between the three groups. Patients in the control group were heavier and had higher body mass index compared to patients with COVID-19 (*p*=0.006; *p*=0.004 respectively). However, there were no differences in these characteristics between control and PASC groups, or COVID-19 and PASC groups. Supplemental figure (Figure S1) graphically depicts patients matching based on height. Median time between diagnosis and CT scan was longer in the PASC (199 days, IQR 126-300 days) compared to the COVID-19 group (44 days, IQR 7-122 days; *p* <0.001).

**Table 1.** Patient demographics. Data are analyzed using Kruskal-Wallis test with Bonferroni correction, and Mann-Whitney U test ^a^. Data are reported as count or median (IQR, interquartile range). p-values are reported for pairwise group comparisons in the following order: controls vs. acute COVID-19; controls vs. long COVID; acute COVID-19 vs. long COVID. p-values: ns, p >0.05; *, p <0.05: **, p <0.01; ***, p <0.001. PASC, post-acute sequelae of COVID-19. BMI, body mass index.

| Variable | Controls | Acute COVID-19 | Long COVID | p-values |
| --- | --- | --- | --- | --- |
| Cohort size, n | 75 | 75 | 75 | .. |
| Male/Female, n | 27/48 | 27/48 | 27/48 | .. |
| Age, years | 53 (36-66) | 55 (43-62) | 50 (37-58) | ns, ns, ns |
| Height, cm | 170 (162-176) | 169 (162.6-177.8) | 170.1 (164.7-176.3) | ns, ns, ns |
| Weight, kg | 89.4 (72.6-101) | 76.4 (65.6-88.6) | 82.5 (66.6-95.6) | **, ns, ns |
| BMI, kg·m<sup>-2</sup> | 30.6 (25.5-34.4) | 26.7 (23.5-30.4) | 28.3 (24.7-32.2) | **, ns, ns |
| Time since diagnosis to CT scan, days<sup>a</sup> | .. | 44 (7-122) | 199 (126-300) | *** |

### Airway luminal area

When the COVID-19 and PASC groups were compared airway luminal area did not differ between these two groups. Among all three study groups difference in trachea average measurements was not significant (p > 0.05). We did not see difference in right main bronchus luminal area between controls and PASC patients. In contrast to that our findings in other bronchi showed significant increase in luminal area in PASC group compared to controls.

For most of the large conducting airways (except trachea and right main bronchus) we observed larger airway luminal areas in COVID-19 group as well as in PASC group compared to controls. Airway cross-sectional area in COVID-19 group was 23.1% (IQR 19.9-37.3%) larger than in control group, and in PASC group 21.6% (IQR 13.5-32.7%) larger compared to controls (difference is calculated as a percent change from healthy controls). Group data are presented in Table 2 with individual data on Figure 3. Findings of additional analysis for large conducting airways middle points are presented in supplemental materials (Table S1).

**Table 2.** Airway luminal cross-sectional areas of the seven main conducting airways in control, acute COVID-19 and long COVID groups. Data are analyzed using Kruskal-Wallis test with Bonferroni correction and reported as median (IQR, interquartile range) of average values measured at proximal, middle, and distal points of each conducting airway. p-values are reported for pairwise group comparisons in the following order: controls vs. acute COVID-19; controls vs. long COVID; acute COVID-19 vs. long COVID. p-values: ns, p >0.05; *, p <0.05: **, p <0.01; ***, p <0.001. PASC, post-acute sequelae of COVID-19.

| Airway luminal area | Controls | COVID-19 | PASC | p-values |
| --- | --- | --- | --- | --- |
| Trachea, mm<sup>2</sup> | 217.7 (185.3-283) | 241.3 (203.3-293.7) | 235.0 (205.0-298.7) | ns, ns, ns |
| Right main bronchus, mm<sup>2</sup> | 149.3 (117.3-186.7) | 179.7 (150.6-212.4) | 167.0 (146-205.3) | *, ns, ns |
| Bronchus intermediate, mm<sup>2</sup> | 83.9 (69.1-102.3) | 104.1 (88.6-126.7) | 96.2 (83.2-118.2) | ***, **, ns |
| Right upper lobe bronchus, mm<sup>2</sup> | 51.1 (45.8-68.5) | 74.3 (59.7-86.6) | 70.8 (60.5-81.6) | ***, ***, ns |
| Left main bronchus, mm<sup>2</sup> | 104.4 (91.5-132.4) | 123.7 (109.7-149.3) | 117.2 (102.7-144) | **, *, ns |
| Left lower lobe bronchus, mm<sup>2</sup> | 43.0 (38.0-57.0) | 57.9 (46.9-71.8) | 52.3 (46.9-65) | **, *, ns |
| Left upper lobe bronchus, mm<sup>2</sup> | 65.2 (51.5-78.3) | 79.6 (66.6-94.2) | 82.7 (65.7-92.9) | **, ***, ns |

**Figure 3.**
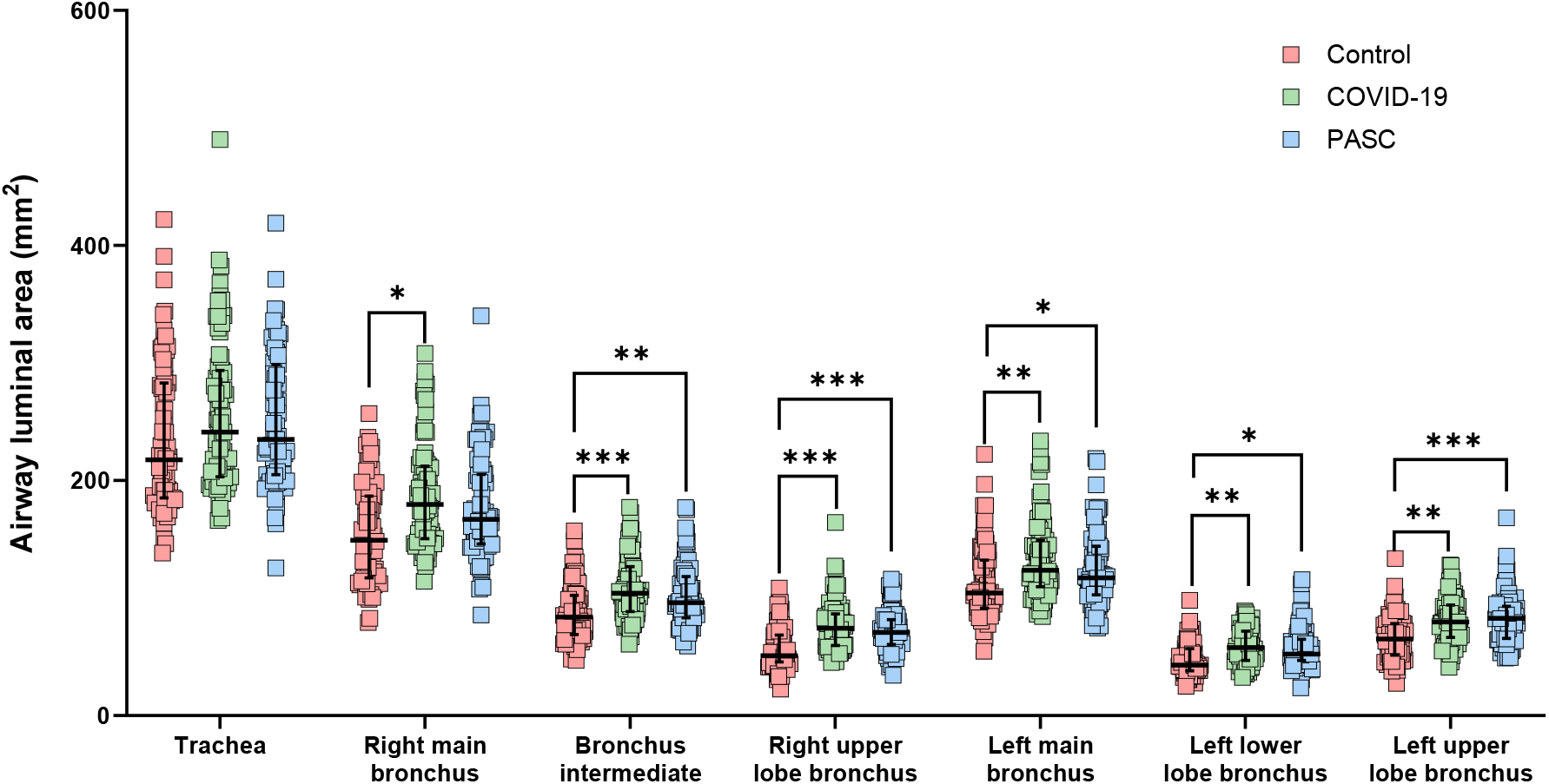
Individual data. Airway luminal cross-sectional areas of the seven main conducting airways in patients with long COVID and acute COVID-19 compared to healthy controls. Data are reported as medians of average values measured at proximal, middle, and distal point of each airway with upper and lower limits and IQR, interquartile range. PASC, post-acute sequelae of COVID-19. *, p <0.05: **, p <0.01; ***, p <0.001. PASC, post-acute sequelae of COVID-19.

## Discussion

### Main findings

The main study findings indicate that patients diagnosed with PASC have larger airway luminal area in most conducting airways compared to healthy controls who have had their imaging performed before the start of the COVID-19 pandemic. Whereas average airway luminal area in the PASC group was not different compared to the COVID-19 group. These findings suggest that SARS-CoV-2 infection might be associated with persistent greater airway luminal area and such association may contribute to respiratory manifestations of both acute COVID-19 and PASC. Additional analysis of asymptomatic patient airways compared to the PASC cohort and healthy controls would have been beneficial in determining if increased airway luminal area is an underlying condition predisposing to infection or consequence of the infection. However, recruitment of such cohort would be difficult as most likely asymptomatic patients with COVID-19 do not have indications for CT imaging.

### Pathophysiology of airway changes

We have yet to understand if increased airway luminal area is an underlying condition in patients with COVID-19 or is a sequela of infection. Inside the new host, SARS-CoV-2 enters respiratory epithelial cells via angiotensin-converting enzyme 2 (ACE2) receptors.^15^ ACE2 receptors are commonly expressed in a variety of organs/tissues, including lungs, and epithelial cells of the trachea and bronchi. Considering that the SARS-CoV-2 uses ACE2 receptors for cell entry, its persistence at the entry site might be a contributing factor of inflammation with consequent bronchial dilation. Alternatively, it is known that patients with irreversible pathological widening of airways, also known as bronchiectasis, have impaired mucociliary clearance. This impairment as an underlying condition may lead to increased frequency of respiratory infection, caused by bacterial and viral agents, presumably including SARS-CoV-2.

Observational studies conducted since the COVID-19 pandemic onset have been focused on analyses of follow-up CT images in patients with previous COVID-19 and showed lung and airway abnormalities.^16-23^ Signs of fibrosis and ground-glass opacifications were seen 3 months after initial symptoms in more than half of the patients with rate of persistent respiratory symptoms in these patients up to ~40%.^16^ Cross-sectional studies report fibrosis of the interstitial lung tissue and bronchiectasis on CT imaging in patients even 6 months after discharge from the hospital.^17,18^ Fibrotic changes on CT were associated with age, longer hospitalization, acute respiratory distress syndrome, and non-invasive mechanical ventilation. Bronchial dilation was thought to be caused by scarred pulmonary tissue traction force and referred to as traction bronchiectasis.^17,18^

The pathogenesis of bronchiectasis as a disease is not fully understood. However, some patients with this condition have a history of preceding infectious or inflammatory insults to the airway while others have underlying condition with mucociliary clearance dysfunction.^24^ Often this dysfunction can be observed in smokers; thus, patients with the history of smoking were excluded from the study cohort. Bronchial dilation and compromised mucociliary clearance lead to mucus plugging and microorganism colonization with activation of host defense inflammatory mechanism to facilitate elimination of foreign particles. Paradoxically, this defense mechanism leads to further damage of bronchial wall and compromises respiratory tract protection.

These aforementioned changes observed via CT imaging have brought our group to conduct our previous study, where findings showed greater airway areas in patients with COVID-19 compared to healthy controls.^8^ But it remained undetermined if the enlarged airway area persists in those with a history of COVID-19 or slowly regresses over time. Studies described in literature are primarily focused on qualitative assessment of airways on chest CT, rather than on direct measurements of airways luminal area size and comparison with a healthy control group. In patients with PASC months after initial COVID-19 infection we see pattern of increased airway size compared to controls. Although, on the additional analysis we see a decrease of airway luminal area at middle points of right and left main bronchi only, in patients with PASC compared to patients with COVID-19 only (Table S1). This finding allows to assume possible regression of changes over time in these two types of bronchi.

### Potential clinical implications

While many studies have addressed changes on CT scans during acute infection or post COVID-19,^16-23^ novelty of our study is in quantitative assessment of airway anatomy by measuring luminal areas with a three-dimensional organ reconstruction software. Our findings are supportive of above-mentioned studies that show bronchial dilation on CT.

Increase in airway luminal area may be considered evidence of a transient condition that subside over time or possible airway remodeling. However, we have not been able to determine whether these changes in airway size are reversible. Possible mechanisms of respiratory symptoms in patients with PASC have been previously described by other studies as air trapping on chest CT imaging, also known as small airway disease.^19,25,26^ Our study findings contribute to this existing knowledge.

### Limitations

Our study has some limitations. First, this retrospective, cross-sectional study focused primarily on luminal area of central conducting airways, thus, limited mechanistic insights may be inferred from our results *per se*. Second, longitudinal data were not available, thus, we could not assess baseline airway anatomy prior to COVID-19 infection or assess follow-up CT in the same cohort of patients. Third, our patient cohorts were not stratified based on the severity of COVID-19 infection in disease groups. Hence, we cannot assess if airway luminal areas are associated with COVID-19 or PASC severity. Fourth, CT images were obtained at end-inspiratory lung volume that was not standardized to total lung capacity. Although, considering that study was evaluating differences in the large airways, effect of lung volume on their diameter would be less likely than in distal bronchi.^27^ Finally, pulmonary function was not systematically assessed in this study.

## Conclusion

These findings suggest that both patients with COVID-19 and PASC have enlarged airway luminal area in the majority of large conducting airways, compared to sex-, age-, and height-matched healthy controls. This study findings contribute to the existing knowledge on PASC pathophysiology as evidence of possible underlying condition or structural airway changes following infection.

## Data availability

Study data cannot be shared publicly because of Institutional Review Board restrictions. Individual participant data underlying the results reported in this publication may be made available to approved investigators for secondary analyses. A scientific committee will review requests for the conduct of protocols approved or determined to be exempt by an Institutional Review Board. Requestors may be required to sign a data use agreement. Data sharing must be compliant with all applicable Mayo Clinic policies.

## Acknowledgements

This research was supported, in part, by National Heart, Lung, and Blood Institute (F32HL154320 to JWS; 5R35HL139854 to MJJ). AHR was supported by a Postdoctoral Fellowship from the Natural Sciences and Engineering Research Council of Canada.

## Author contributions

S.Z. and A.H.R. have assessed and verified the underlying data reported in the manuscript. Study conception and design: S.Z., A.J.M., J.W.S., C.C.W., A.H.R. Acquisition, analysis, or interpretation of data: S.Z., A.J.M., E.A.O., A.H.R. Drafting of the manuscript: S.Z., C.C.W., J.W.S., A.H.R. Administrative, technical, or material support: P.B.D., R.G., R.T.H., B.J.B., B.T.W., J.G.R., M.J.J. All authors contributed to revising the manuscript, and all authors approved the final version of the manuscript.

