## Supplementary material for "Increased luminal area of large conducting airways in patients with COVID-19 and post-acute sequelae of COVID-19 A retrospective case-control study"

This material has been provided to give readers additional information about the work.

**Most recent update:** February 22, 2024

### Increased luminal area of large conducting airways in patients with COVID-19 and post-acute sequelae of COVID-19
### A retrospective case-control study

#### Supplementary material

| Airway luminal area | Controls | COVID-19 | PASC | p-values |
|---|---|---|---|---|
| Trachea, mm2 | 209 (183-280) | 240 (198-288) | 227 (193-295) | ns, ns, ns |
| Right main bronchus, mm2 | 143 (118-179) | 179 (151-208) | 155 (137-192.5) | **, ns, * |
| Bronchus intermediate, $mm^2$ | 79.1 (60.9-95.9) | 98.7 (85.8-116) | 88 (78.6-112) | ***, ns, ns |
| Right upper lobe bronchus, $mm^2$ | 52.2 (44.4-67.2) | 69.7 (55.5-83.4) | 61.1 (53.8-75.3) | ***, *, ns |
| Left main bronchus, $mm^2$ | 87.3 (75.5-115) | 120 (97-143) | 99.8 (87.2-124.8) | ***, ns, * |
| Left lower lobe bronchus, $mm^2$ | 42.6 (38-56.1) | 51.3 (39.5-63.7) | 48.9 (39-58.8) | ns, ns, ns |
| Left upper lobe bronchus, $mm^2$ | 63.1 (50.3-77.5) | 78.7 (65.7-90.7) | 81.3 (64.3-93.4) | **, ***, ns |

**Supplemental Table S1.** Airway luminal cross-sectional areas of the seven main conducting airways in control, acute COVID-19 and long COVID groups. Data are analyzed using Kruskal-Wallis test with Bonferroni correction and reported as median (IQR, interquartile range) of airway luminal areas measured at midpoints of each airway. p-values are reported for pairwise group comparisons in the following order: controls vs. acute COVID-19; controls vs. long COVID; acute COVID-19 vs. long COVID. p-values: ns, p >0.05; *, p <0.05: **, p <0.01; ***, p <0.001. PASC, post-acute sequelae of COVID-19.

#### Supplemental notes

This additional analysis results are overall similar to the main study findings. There were no differences in trachea area between groups. There are some statistical discrepancies that show decrease in midpoint areas of the right and left main bronchi in PASC group compared to COVID-19 group, but do not show any differences between control and PASC group. The direction of changes is identical to the main analysis of average areas and indicates greater airways area in large conducting airways in COVID-19 and PASC groups compared to controls, where the difference is significant.

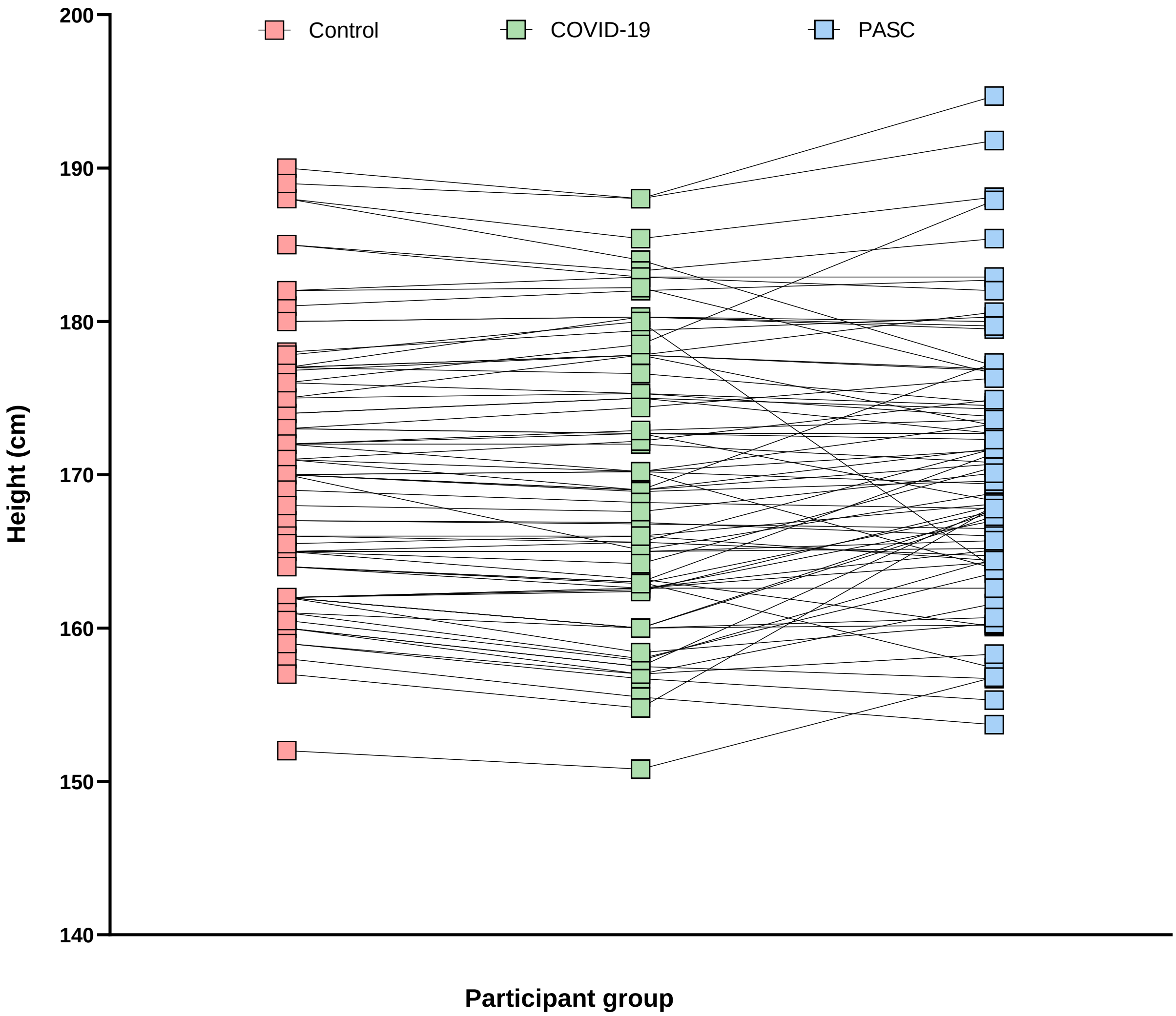

**Supplemental Figure S1.** Participant matching algorithm based on height of control, acute COVID-19 and long COVID group. Height of each participant is represented by symbols; matched pairs are connected by lines. PASC, post-acute sequelae of COVID-19.

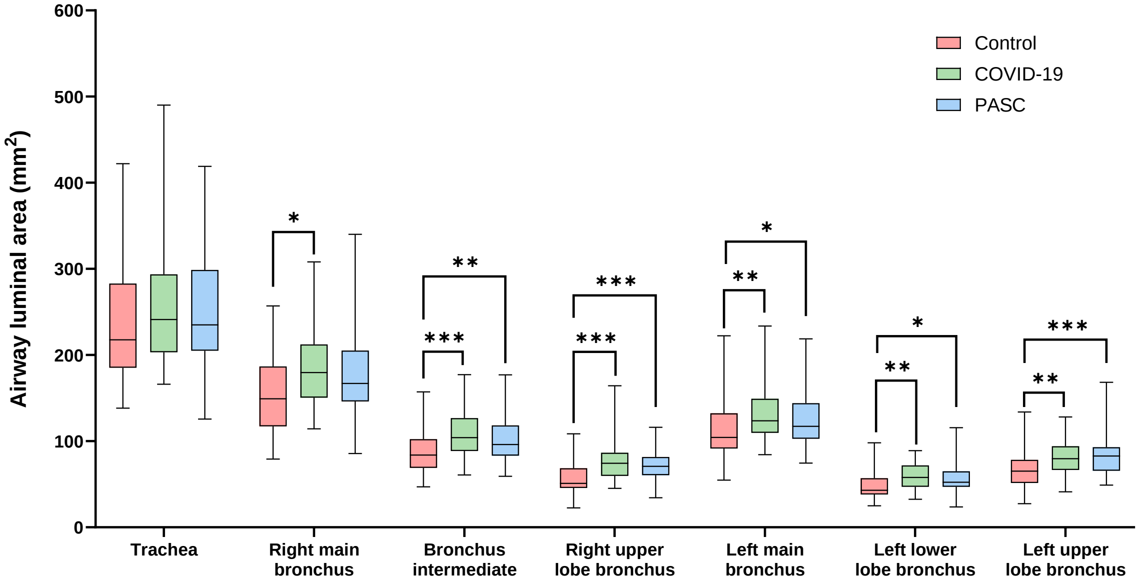

**Supplemental Figure S2.** Airway luminal cross-sectional areas of the seven main conducting airways in patients with long COVID and acute COVID-19 compared to healthy controls. Data are reported as medians of average values measured at proximal, middle, and distal point of each airway with upper and lower limits and IQR, interquartile range. PASC, post-acute sequelae of COVID-19. p-values: *, p <0.05: **, p <0.01; ***, p <0.001.
